# Undetected isoniazid resistance leads to rifampicin-resistant tuberculosis: a prospective observational study and transmission modelling analysis

**DOI:** 10.64898/2026.05.22.26353840

**Authors:** Ruan Spies, Nguyen Hong Hanh, Phan Trieu Phu, Luong Kim Lan, Kim Lan, Ngo Ngoc Hue, Nguyen Le Quang, Do Dang Anh Thu, Nguyen Thi Le Huong, Tran Le Thi Ngoc Thao, Trinh Thi Bich Tram, Vu Thi Ngoc Ha, Dang Thi Minh Ha, Nguyen Phuc Hai, Nguyen Hung Thuan, Tran Thi Kim Quy, Nguyen Huu Lan, Viola Dreyer, Stefan Niemann, Derrick Crook, Le Hong Van, Guy E. Thwaites, Nguyen Thuy Thuong Thuong, Marc Choisy, James A. Watson, Timothy M. Walker

## Abstract

**Background:** Isoniazid resistance is the most common form of drug-resistant tuberculosis (TB) globally. However, WHO-recommended molecular tests available to most TB patients worldwide detect rifampicin resistance only, risking under-treatment of isoniazid-resistant, rifampicin-susceptible TB (HR-TB) and the subsequent emergence of rifampicin resistance.

**Methods:** This prospective study, conducted between March 2020 and July 20204, aimed to collect and archive sputum specimens from all adults diagnosed with rifampicin-susceptible pulmonary TB in Ho Chi Minh City, Vietnam. Cases were participants who developed rifampicin-resistant recurrence; controls had rifampicin-susceptible recurrence or no recurrence. Whole-genome sequencing of paired isolates distinguished acquired rifampicin resistance from reinfection. The effect of pre-existing isoniazid resistance on rifampicin resistance acquisition was estimated using inverse probability of treatment weighting, and the projected epidemiological impact of routine HR-TB testing on the incidence of rifampicin-resistant TB (RR-TB) was modelled.

**Findings:** 42,843 people were diagnosed with TB during the study period, from whom we archived 33,843 sputum samples. We enrolled 1,241 participants, 873 (70·4%) of whom had analysable data. 51/873 (5·8%) acquired rifampicin resistance, of whom 49/51 (96·1%) had isoniazid resistance at their index TB episode that was not detected by routine diagnostics. The weighted risk of acquired rifampicin resistance was 2·98% (95% CI 2·08-4·50) in participants with programmatically undetected isoniazid resistance, versus 0·03% (0·00-0·08) in participants with isoniazid-susceptible TB (risk ratio 105·42 (33·43-309·69)). Modelling projected that universal HR-TB diagnosis and treatment would reduce RR-TB incidence by 46% (35-61) over 10 years in Vietnam, with reductions of 26% (12-43) projected even where HR-TB prevalence was as low as 5%.

**Interpretation:** Undetected, under-treated HR-TB confers a 100 fold increased risk of acquiring rifampicin resistance. Routine isoniazid susceptibility testing combined with effective HR-TB treatment could substantially reduce the burden of RR-TB.

**Funding:** Wellcome

## Research in context

### Evidence before this study

We searched PubMed from database inception to January 2026 for studies investigating the risk of acquired rifampicin resistance among patients with isoniazid-resistant, rifampicin-susceptible tuberculosis (HR-TB), using the terms “tuberculosis”, “isoniazid resistance”, “rifampicin resistance”, “multidrug resistance”, “acquired resistance”, “treatment outcome”, and “whole-genome sequencing”. The largest body of evidence comes from a systematic review and meta-analysis (Gegia et al., *Lancet Infect Dis* 2017) that pooled 19 cohort studies and 33 randomised trials encompassing 3,744 patients with HR-TB and 19,012 with drug-susceptible disease. Treatment with the WHO-recommended regimen for new patients resulted in failure in 11% (95% CI 6–17), relapse in 10% (5–15), and acquired multidrug resistance in 8% (3–13) of patients with HR-TB, compared with 2% (1–3), 5% (2–7), and 1% (0–2) in those with drug-susceptible disease. However, the constituent studies were conducted predominantly between the 1970s and early 2000s, when programmatic treatment success rates were substantially lower than today, and none used whole-genome sequencing (WGS) to distinguish true *de novo* resistance acquisition from reinfection with an already-resistant strain, meaning the reported effect sizes conflate two mechanistically distinct processes. A single longitudinal cohort study from Ho Chi Minh City, Vietnam (Srinivasan et al., *Clin Infect Dis* 2020) used WGS in 239 patients with HR-TB and found that *de novo* MDR-TB emergence (6/239, 2·5%) and reinfection with a different MDR-TB strain contributed equally to recurrent MDR-TB, but this study enrolled only HR-TB patients without an isoniazid-susceptible comparator group, precluding estimation of relative risk or population attributable fraction.

### Added value of this study

We assembled the largest cohort to date of people with recurrent tuberculosis with paired whole-genome sequencing data at both the index and recurrent episodes, drawn from a city-wide sampling frame covering all approximately 40,000 adults notified with rifampicin-susceptible TB in Ho Chi Minh City over four years. By using WGS to classify each recurrence as relapse or reinfection and applying inverse probability of treatment weighting within a population-weighted design, we show that undetected isoniazid resistance confers a 100-fold increased risk of acquiring rifampicin resistance (risk ratio 105, 95% CI 33–310), an effect far larger than meta-analytic estimates that could not account for reinfection misclassification. We further demonstrate that concurrent pyrazinamide or ethambutol resistance compounds this risk substantially. Using a deterministic two-strain transmission model calibrated to Vietnamese epidemiological data, we project that universal HR-TB detection and treatment could reduce RR-TB incidence by 46% (35–61) over 10 years, with meaningful reductions (26%, 12–43) projected even where HR-TB prevalence is as low as 5%.

### Implications of all the available evidence

Pre-existing isoniazid resistance is a far more important driver of acquired rifampicin resistance than previously recognised. Once reinfection is excluded, the risk among patients with undetected HR-TB treated with first-line therapy is orders of magnitude greater than in those with isoniazid-susceptible disease. These findings suggest an ethical imperative to offer rapid isoniazid resistance testing at tuberculosis diagnosis wherever cost-effectiveness allows. This is now technically feasible as assays combining *M. tuberculosis* detection with simultaneous isoniazid and rifampicin resistance testing on existing low-complexity platforms have recently been developed for near-point-of-care use. Paired with effective HR-TB treatment, routine isoniazid susceptibility testing could substantially reduce the global burden of RR-TB.

Tuberculosis (TB) kills more people each year than any other pathogen, with an estimated 1·23 million deaths in 2024.^1^ Rifampicin-resistant TB (RR-TB) remains a particular concern as it is harder to treat, with worse outcomes than rifampicin-susceptible disease.^1^

The World Health Organization (WHO) recommended the first rapid, low-complexity molecular test for the detection of *Mycobacterium tuberculosis* complex in 2011.^2^ Xpert MTB/RIF was the first in class, but as with platforms that followed, the detection of rifampicin resistance was prioritised, with detection of resistance to other drugs performed through additional reflex-testing, if at all. Isoniazid-resistant, rifampicin-susceptible TB (HR-TB) is the most prevalent form of drug-resistant TB globally but is not detected by Xpert.^3^ Evidence suggesting that isoniazid resistance typically emerges prior to rifampicin resistance has since raised concern that the under-detection of HR-TB, and its consequent treatment with the isoniazid-containing first-line regimen, could select for the emergence of rifampicin resistance.^4,5^

Although a number of studies have demonstrated that transmission of RR-TB, rather than its *de novo* emergence, is the main contributor to RR-TB burden, our recent work from Vietnam estimated that 13-28% of RR-TB may be selected for during treatment.^6,7^ Previous studies have reported a strong association between HR-TB and subsequent RR-TB; however, because they were unable to distinguish reinfection with a different strain from true acquisition of rifampicin resistance in the original infecting strain, they may have underestimated the effect of HR-TB on acquired RR-TB.^5^ Here, we use whole-genome sequencing (WGS) to quantify the risk of acquiring rifampicin resistance due to undetected HR-TB, and model the population-level impact of improved HR-TB detection and treatment on the incidence of RR-TB.

## Methods

### Study setting and design

Ho Chi Minh City, Vietnam’s largest city, records more than 10,000 TB diagnoses annually and has the highest burden of TB and RR-TB in the country.^8^ Outpatient TB care is delivered through a network of 23 District TB Units (DTUs), although all patients diagnosed with RR-TB are referred to the central Phạm Ngọc Thạch Hospital for treatment initiation. Under the National TB Programme (NTP), rifampicin-susceptible TB (RS-TB) is treated with 8 weeks of rifampicin, isoniazid, pyrazinamide and ethambutol, followed by 16 weeks of daily rifampicin isoniazid, and ethambutol. Ethambutol is continued throughout to prevent acquisition of rifampicin resistance in case of undiagnosed HR-TB, which accounts for approximately 20% of all TB in Vietnam.^9,10^

We conducted a prospective observational study among all adults diagnosed with Xpert-confirmed pulmonary RS-TB in Ho Chi Minh City between March 2020 and July 2024. We aimed to collect and archive sputum samples from all patients diagnosed with RS-TB. Participants were then recruited into three sub-cohorts depending on their outcome: no recurrence, recurrence with RS-TB, or recurrence with RR-TB. The study was originally designed as two nested case-control studies comparing isoniazid resistance prevalence between outcome groups **(Supplementary methods)**. Linkage to NTP notification data, combined with efforts to identify every patient with recurrent TB across the four-year study period, subsequently yielded population-level denominators for each outcome stratum, enabling a weighted analysis from which absolute risks could be derived.

This study received ethical approval from the Institutional Review Board at Phạm Ngọc Thạch Hospital (643/PNT-HDDD) and the Oxford Tropical Research Ethics Committee (reference no. 51–19). Written informed consent was obtained from all participants prior to enrolment. Participant identifiers (name, date of birth, address) were retained in a secure, access-restricted database for the purposes of specimen linkage; all analysis was conducted using de-identified study ID numbers, with identifiers accessible only to designated study staff.

### Study procedures

TB recurrence was defined as Xpert-confirmed TB occurring at least 5 months after the index episode, irrespective of treatment status, thereby capturing treatment failure, post-treatment relapse, and reinfection. Follow-up duration varied between participants and was determined by the interval from index diagnosis to recurrence or the end of the study period (no recurrence). Patients with a potential TB recurrence were identified by clinical staff across DTUs and Phạm Ngọc Thạch Hospital. The study team was then notified and matched the incoming case to archived index specimens using name, year of birth and address. Eligible participants were enrolled and a study specimen collected directly by the study team. Given the reliance on name, year of birth and address for linkage, all candidate matches were manually reviewed by study staff to confirm identity before enrolment, reducing the risk of misidentification from duplicate or similar names.

Participants with recurrence were enrolled consecutively as identified across all DTUs and Phạm Ngọc Thạch Hospital throughout the study period. Participants without recurrence were randomly selected from the same DTU as a corresponding RS-TB participant with recurrence, matched by sex and age (±10 years).

All participants provided a baseline sputum specimen for culture and WGS, and those with recurrence an additional sample upon diagnosis of their second episode. All study specimens were transported to and processed at a single central research laboratory. Sputum decontamination, processing, culture, and WGS have been described previously.^7,11^ Species and lineage identification, drug susceptibility prediction, and pairwise single nucleotide polymorphism (SNP) differences were obtained using the GPAS *Mycobacterium* pipeline v2.2.1.^12^ All participants were managed by the NTP throughout the study. WGS was performed in batches after study completion and results did not influence clinical management.

### Study variables

Cases were defined as participants with a recurrent TB episode in which rifampicin resistance was detected by WGS and the recurrence isolate was consistent with relapse rather than reinfection (≤12 SNP differences between index and recurrence isolates).^13^ Controls were participants with no recurrence, rifampicin-susceptible recurrence, or rifampicin-resistant recurrence attributable to reinfection (>12 SNP differences). As a sensitivity analysis, case and control classification was repeated using a more conservative ≤5 SNP threshold to define same-strain recurrence. The primary exposure was pre-existing isoniazid resistance, defined as any isoniazid resistance-conferring mutation in the index isolate according to the WHO mutation catalogue.^14^ Pre-existing pyrazinamide and ethambutol resistance were recorded as secondary exposures to evaluate whether concurrent companion drug resistance further increased the risk of rifampicin resistance acquisition among participants with HR-TB.

### Statistical analysis

The primary analysis estimated the marginal effect of pre-existing isoniazid resistance on the acquisition of rifampicin resistance. Because outcome-stratified sampling over-represents rarer outcomes relative to the source population, sampling weights were derived from population-to-sample ratios for each sub-cohort based on NTP notification data **(Supplementary Methods)**. To adjust for confounding, propensity scores for baseline isoniazid susceptibility were estimated using logistic regression including age, sex, diabetes, HIV status, and *M. tuberculosis* lineage, and inverse probability of treatment weights were calculated and stabilised by truncation at the 1st and 99th percentiles. Covariate balance between isoniazid-resistant and isoniazid-susceptible groups before and after weighting was assessed using absolute standardised mean differences (SMD), with adequate balance defined as all SMDs below 0·1.

Final analysis weights were the product of sampling and propensity score weights. Weighted risks of acquired RR-TB were estimated for isoniazid-resistant and isoniazid-susceptible groups, from which the adjusted risk ratio was derived. The population attributable fraction (PAF) was estimated using Miettinen’s formula.^15^ Uncertainty was estimated by non-parametric bootstrap resampling (1,000 iterations), with all weighting procedures repeated within each resample.

To investigate whether concurrent resistance to companion drugs further increased the risk of rifampicin resistance acquisition among participants with isoniazid- resistant TB, we compared the weighted risk of acquired rifampicin resistance between those with isoniazid monoresistance and those with concurrent pyrazinamide or ethambutol resistance, using sampling weights only.

We adapted a previously published deterministic compartmental model of TB transmission in Vietnam^16^ to incorporate two strains (rifampicin-susceptible and rifampicin-resistant) and estimate the population-level impact of diagnosing and treating HR-TB on the incidence of RR-TB. TB natural history was represented by six states per strain: susceptible, infection, non-infectious disease, asymptomatic and symptomatic disease, and post-treatment, with bidirectional transitions between non-infectious, asymptomatic, and symptomatic states reflecting the dynamic progression and regression of untreated TB.^17^ Acquired rifampicin resistance was modelled as arising through treatment failure and post-treatment relapse. The model was calibrated to WHO estimates of TB and RR-TB incidence in Vietnam (2000–2024)^1^ and to the empirical proportion of RR-TB attributable to acquired resistance (13–28%)^7^ using history matching with emulation.^18^ Full model structure, equations, and parameterisation are detailed in the **Supplementary Methods**.

We simulated scaling up HR-TB diagnosis and appropriate treatment from 2025 at four coverage levels (20%, 50%, 80%, and 100%), compared with a baseline of 0%. Under intervention scenarios, the probability of acquiring rifampicin resistance depended on diagnostic coverage, treatment efficacy of the WHO-recommended HR-TB regimen (estimated at 89%, the current first-line success rate in Vietnam)^19–21^ and the empirically estimated PAF. Outcomes were projected RR-TB incidence and relative reduction in RR-TB incidence under intervention scenarios by 2035. To assess the robustness of our findings, we recalculated the PAF, using Levin’s formula^22^, across nine scenarios defined by three levels of HR-TB prevalence (5%, 10%, 20%) and three relative risks for acquired rifampicin resistance (the empirically observed estimate and 2-fold and 5-fold attenuations thereof), and repeated the universal coverage intervention under each scenario **(Supplementary Methods)**.

## Results

Between March 2020 and July 2024, 42,843 patients were diagnosed with RS-TB in Ho Chi Minh City, from whom we collected and archived 33,843 sputum samples. From this source population, we enrolled 1,241 participants across no-recurrence (n=515), RS-TB recurrence (n=515), and RR-TB recurrence (n=211) sub-cohorts **(Figure 1)**. Median time to RS-TB recurrence was 12·3 months (9·6-18·1) and 9 months (6-14·8) for RR-TB recurrence; median follow-up of participants without recurrence was 8·6 months (IQR 5·9-20·7). Extended follow-up through NTP records to June 2025, providing at least 12 additional months of observation for all no-recurrence participants, identified one additional RR-TB and three RS-TB recurrences in this group. Participants were excluded for inability to collect sputum (n=25 [2%]), non-viable culture or DNA extraction (n=251 [20%]), non-tuberculous mycobacteria (n=34 [3%]) and WGS-confirmed rifampicin resistance at the index episode (n=54 [4%]), leaving 873 (70%) participants with analysable data **(Figures 1** and **S3).** Among the 54 excluded rifampicin-resistant index isolates, 14 (26%) harboured *rpoB* mutations outside the rifampicin resistance-determining region (V170F [n=5] or I491F [n=9]) that are undetectable by Xpert MTB/RIF.

**figure 1:**
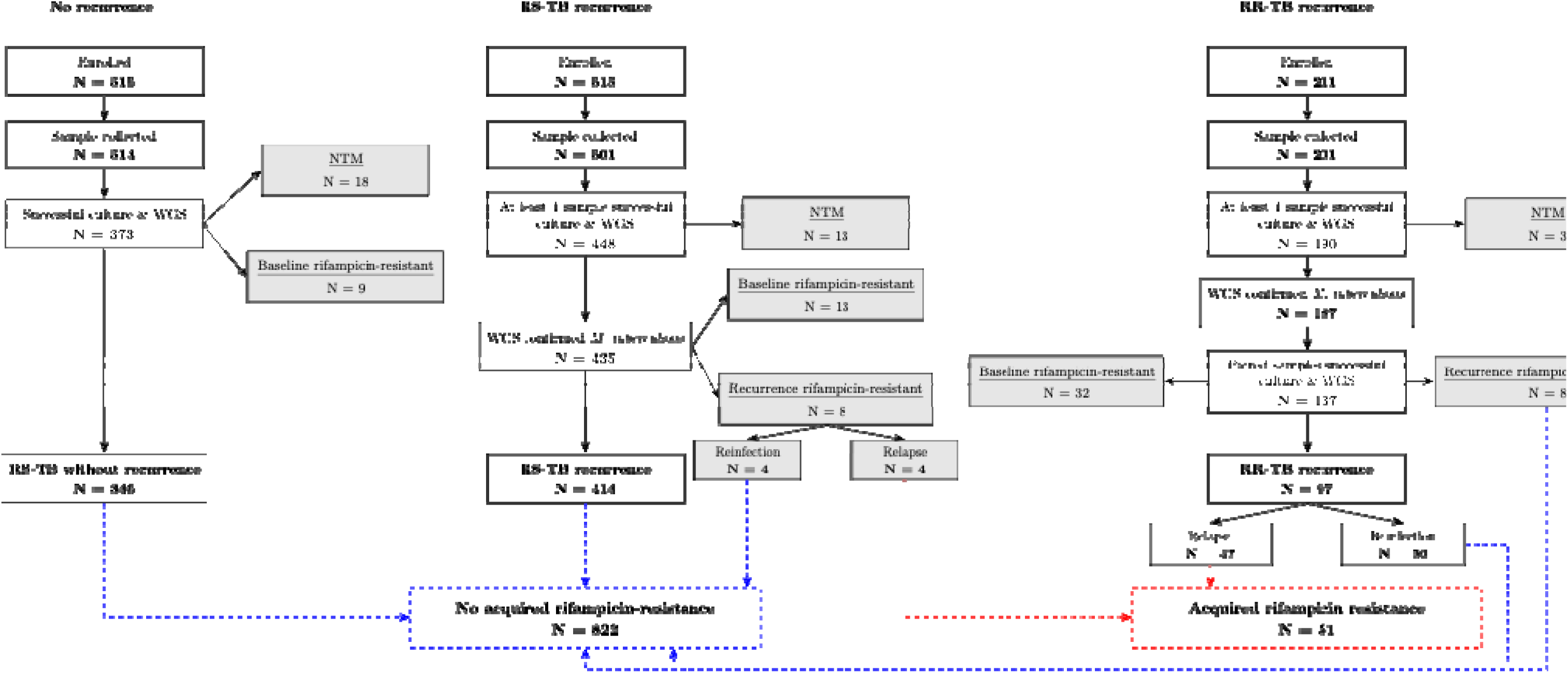
Study flow diagram. participants were enrolled into three sub-cohorts defined by recurrence and rifampicin susceptibility status on Xpert MTB/RIF. Exclusions at each stage are show culture failure, non-tuberculous mycobacteria (NTM), and WGS-confirmed rifampicin resistance at the index episode. For participants with paired index and recurrence isolates, WGS-derived pairwise SNP distances were used to classify recurrences as relapse (≤12 SNPs) or reinfection (>12 SNPs). Acquired rifampicin resistance defined as relapse with a new rifampicin resistance-conferring mutation in the recurrence isolate. Blue dashed lines indicate participants classified as having no resistance; red dashed lines indicate participants classified as having acquired rifampicin resistance. See **Figure S3** for further detail on exclusions.

Among participants with paired isolates, WGS reclassified over half (50/97) of RR-TB recurrences as reinfections with a different RR-TB strain rather than true acquired resistance. Pooling across sub-cohorts, 51 participants were classified as having acquired rifampicin resistance and 822 as having no evidence of acquired resistance **(Figure 1)**. Using the more conservative ≤5 SNP threshold reclassified only one participant from case to control, indicating case ascertainment was insensitive to this choice. Most acquired rifampicin resistance occurred in participants classified as relapse (n=37, 73%) while 14 (27%) cases occurred during treatment failure.

Participants who acquired rifampicin resistance were less commonly male (67% vs 81%, p=0·03), more frequently diabetic (49% vs 32%, p=0·01), and their index isolates were more commonly Lineage 2 (92% vs 66%, p<0·001) compared with those who did not **(Table 1)**. Pre-existing resistance to isoniazid (96% vs 18%), pyrazinamide (33% vs 3%), and ethambutol (26% vs 1%) were associated with acquired rifampicin resistance (all p<0·001).

**Table 1:** Participant characteristics stratified by outcome status.

|  | No acquired rifampicin<br>resistance<br>(N=822) | Acquired rifampicin<br>resistance<br>(N=51) |
| --- | --- | --- |
| Age, median (IQR) | 53 (45-60) | 52 (39-57) |
| Male sex, n | 663 (81%) | 34 (67%) |
| Diabetes, n | 259 (32%) | 25 (49%) |
| HIV positive, n | 10 (1%) | 2 (4%) |
| *M.tuberculosis* lineage, n |  |  |
| Lineage 1 | 169 (21%) | 3 (6%) |
| Lineage 2 | 543 (66%) | 47 (92%) |
| Lineage 4 | 46 (6%) | 0 (0%) |
| Mixed or unknown | 64 (8%) | 1 (2%) |
| Pre-existing isoniazid resistance, n | 151 (18%) | 49 (96%) |
| Pre-existing pyrazinamide resistance, n | 23 (3%) | 17 (33%) |
| Pre-existing ethambutol resistance, n | 11 (1%) | 13 (26%) |

Among participants with baseline isoniazid resistance, *katG* S315T was the predominant mechanism, identified in 47/49 (96%) of those who acquired rifampicin resistance and 118/151 (78%) of those who did not **(Table S1)**. Concurrent resistance to pyrazinamide or ethambutol was present in 52 of 200 (26%) participants with HR-TB and was more common among those with RR-TB recurrence (30 [47%]) than RS-TB recurrence (15 [19%]) or no recurrence (7 [13%]). The most commonly acquired rifampicin resistance mutation was *rpoB* H445R (n=13, 25%; **Table S2**).

After weighting for sampling probability and baseline confounding **(Figure S1)**, the risk of acquired rifampicin resistance was 2·98% (2·08-4·50) among participants with pre-existing isoniazid resistance, compared to 0·03% (0·00-0·08) among those with isoniazid-susceptible TB, corresponding to a risk ratio of 105·42 (33·43–309·69). The PAF was 95% (87-98), suggesting nearly all acquired rifampicin resistance in this population arose from pre-existing isoniazid resistance treated with first-line therapy under current programmatic conditions. We observed no difference in same-strain RS-TB recurrence risk by isoniazid resistance status, though the study was not powered to detect modest differences in this outcome **(Supplementary results)**.

Among participants with HR-TB, the risk of acquiring rifampicin resistance was 1·98% (1·23–2·94) with isoniazid monoresistance, rising to 13·10% (6·95–31·26) when pyrazinamide or ethambutol resistance was also present. Sample sizes were insufficient to reliably estimate the separate effects of pyrazinamide and ethambutol resistance individually **(Table S3)**.

Without intervention, RR-TB incidence in Vietnam in 2035 was projected to be 7·0 (95% UI: 4·8-11·4) per 100,000 per year, with 15·7% (10·0-24·7) arising from acquired resistance. Under a scenario of universal isoniazid susceptibility testing and appropriate treatment of HR-TB, RR-TB incidence was projected to fall to 3·7 (2·3-6·6) per 100,000 per year by 2035 with 5·5% (2·7–11·6) due to acquired resistance **(Figure 2A)**. The effect was dependent on coverage of diagnostic testing: by 2035, coverage of 20%, 50%, 80%, and 100% reduced RR-TB incidence by 9%, 23%, 37%, and 46%, respectively **(Figure 2B)**. In sensitivity analyses varying isoniazid resistance prevalence (5–20%) and the relative risk for acquired rifampicin resistance (observed, 105 [33–310], 2-fold and 5-fold attenuations) to approximate other settings, universal coverage reduced RR-TB incidence by 26-47% across all scenarios **(Figure 3)**.

**Figure 2:**
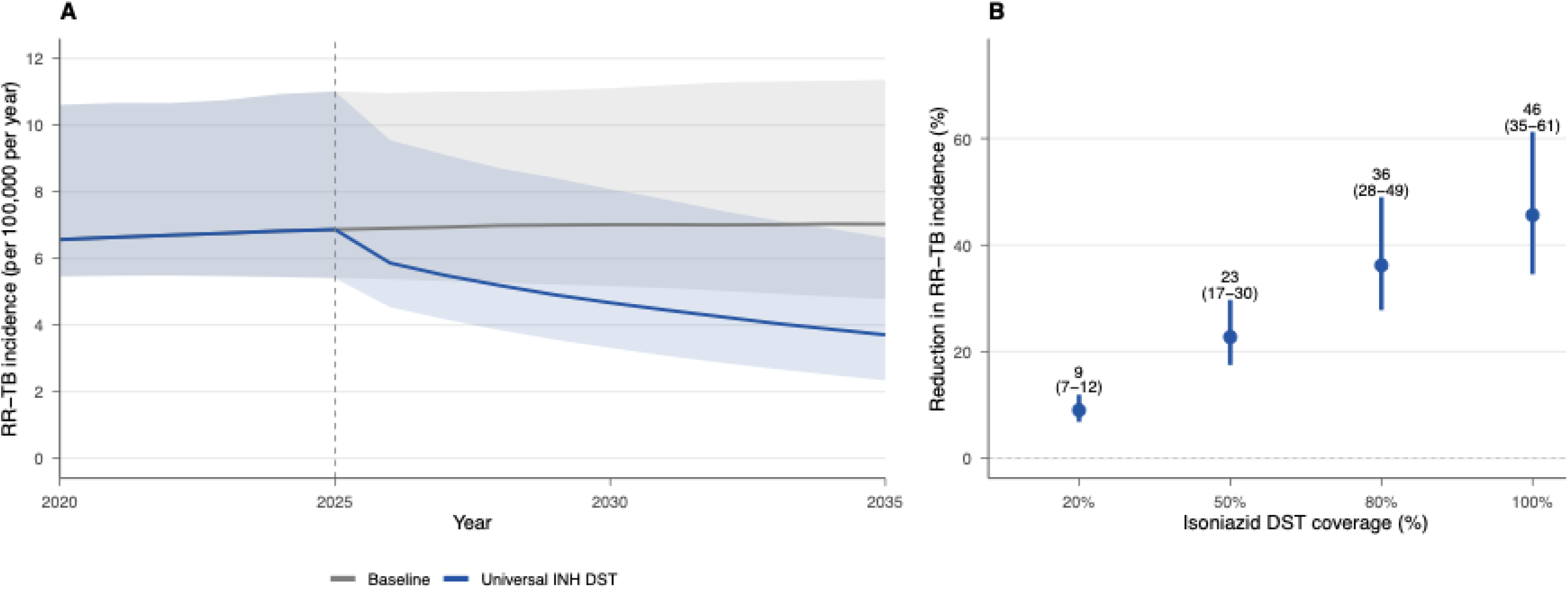
Reduction in rifampicin-resistant tuberculosis incidence in Vietnam under isoniazid drug susceptibility testing scenarios Panel A: Projected RR-TB incidence in Viet Nam under baseline scenario (no systematic isoniazid susceptibility testing) and universal isoniazid susceptibi testing from 2025. Solid lines represent medians and shaded areas 95% uncertainty intervals. Dashed link represents intervention start at 01/01/2025 **Pan** Projected relative reduction in RR-TB incidence in Viet Nam at four levels of isoniazid drug susceptibility testing coverage compared to baseline.

**Figure 3:**
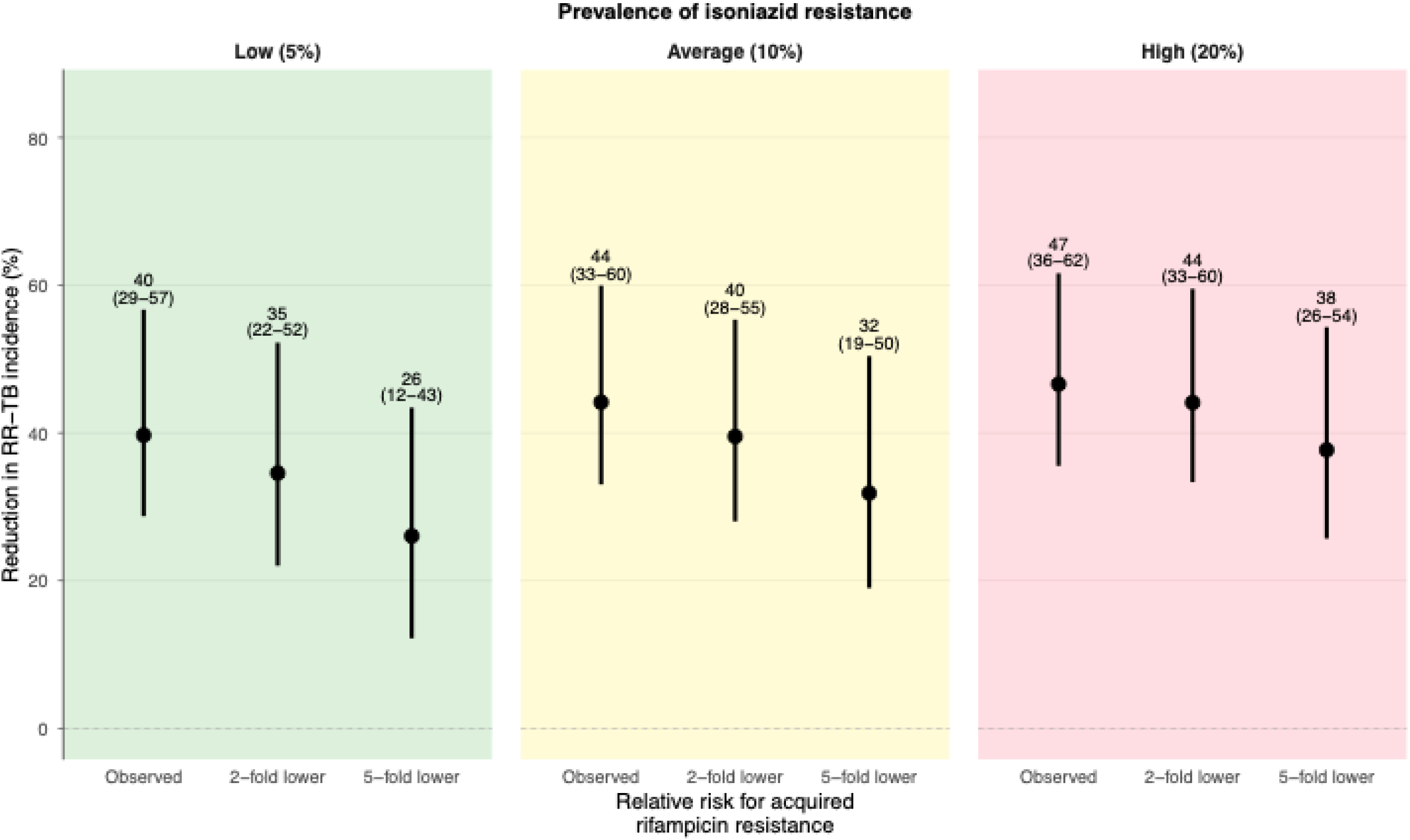
Projected reduction in RR-TB incidence under universal isoniazid susceptibility testing across settings with varying isoniazid resistance prevalence and relative risk for acquired rifampicin resistance. Nine scenarios were constructed by combining three levels of isoniazid resistance prevalence among TB patients (low: 5%, average: 10%, high: 20%) with three levels of the relative risk for acquired rifampicin resistance given pre-existing isoniazid resistance (Observed, 2-fold lower and 5-fold lower). For each scenario, the population attributable fraction was recalculated and the universal coverage intervention re-run across all calibrated parameter sets. Points represent median projected reduction in RR-TB incidence by 2035; horizontal bars represent 95% uncertainty intervals.

## Discussion

In this study we show undetected HR-TB treated with isoniazid-containing first-line therapy increases the risk of acquired rifampicin resistance by over 100-fold. That undetected isoniazid resistance confers an increase in risk was anticipated; the magnitude of this effect was not. Consequently, transmission modelling projected that universal diagnosis and treatment of HR-TB initiated in 2025 could reduce RR-TB incidence in Vietnam by nearly half by 2035. Substantial reductions were projected in other settings even under conservative assumptions about isoniazid resistance prevalence and effect size, suggesting these findings are relevant beyond Vietnam to anywhere HR-TB is not routinely detected.

A previous meta-analysis demonstrated that people with HR-TB treated with first-line therapy acquired multidrug resistance at a 25-fold higher rate than those with drug-susceptible disease.^5^ We observed a far greater effect as WGS allowed us to distinguish true acquired resistance from reinfection with already-resistant strains, a distinction not possible in prior studies. Indeed, over half of programmatically classified RR-TB recurrences in our cohort proved to be reinfections. By minimising outcome misclassification and adjusting for confounders including *M. tuberculosis* lineage^23^, we demonstrate that undiagnosed isoniazid resistance is a much more important cause of acquired rifampicin resistance than previously understood.

Genomic analyses of globally diverse *M. tuberculosis* complex strains have consistently shown that rifampicin resistance emerges on a phylogenetic background of pre-existing isoniazid resistance.^4,24^ Our findings demonstrate that this population-level evolutionary pattern is reflected within individuals, and that the magnitude of the effect is far greater than previously anticipated with nearly all acquired rifampicin resistance attributable to pre-existing isoniazid resistance. In patients with HR-TB, first-line therapy is functionally reduced to three effective drugs during the intensive phase, and to rifampicin monotherapy (or with ethambutol) during the continuation phase. It has previously been argued that isoniazid contributes primarily to the initial bactericidal kill and has little sterilising role thereafter, predicting that its loss should matter most early in treatment.^25^ Although the impact of pyrazinamide resistance supports this (used only for 8 weeks), three quarters of acquired rifampicin resistance in our study was detected as relapse rather than treatment failure, possibly suggesting the critical period for resistance selection extends beyond the intensive phase. Rifampicin monotherapy can select for resistant subpopulations within weeks, which may persist below the threshold of clinical detection and manifest only as relapse after treatment completion.^26^ The dose-response gradient we observed suggests ethambutol has some mitigating effect, and that the risk of acquiring rifampicin resistance may be even greater in countries where this is not used in the continuation phase.

Although only 13–28% of RR-TB in Ho Chi Minh City arises through acquired resistance^7^, the projected reduction in RR-TB incidence under intervention scenarios substantially exceeds this proportion because each case prevented also eliminates a potential source of onward transmission. However, realising this benefit requires diagnostics that detect isoniazid resistance at treatment initiation. Current algorithms relying on Xpert MTB/RIF (or Ultra) alone cannot do this. Alternative assays capable of detecting isoniazid resistance, including Xpert MTB/XDR, moderate-complexity nucleic acid amplification tests, and targeted next-generation sequencing, have become increasingly available in recent years.^27,28^ However, cost and infrastructure constraints have limited their use primarily to reflex testing of rifampicin-resistant isolates, rather than adoption as universal diagnostics.^29^ A recently described assay combining *M. tuberculosis* complex detection with simultaneous rifampicin (including *rpoB* I49F) and isoniazid resistance testing on existing low-complexity GeneXpert platforms suggests near-point-of-care dual susceptibility testing is technically feasible.^30^ Whether isoniazid resistance testing should be deployed universally or targeted to higher-risk subgroups, such as people with previous TB, is a question of cost-effectiveness not addressed by our study. However, the magnitude of the effect we estimate suggests such an analysis is a priority.

Improved diagnosis will only translate into fewer acquired rifampicin-resistance cases if it is paired with effective treatment. WHO recommends the addition of levofloxacin to rifampicin, pyrazinamide and ethambutol therapy for HR-TB, but this carries very low certainty of evidence based on predominantly observational studies.^19,20^ The on-going FLIRT trial (ACTRN12624000654550), a phase III, randomised controlled trial evaluating levofloxacin-based therapy for HR-TB should provide evidence to guide treatment when it reports. The WHO-recommended HR-TB regimen assumes pyrazinamide and ethambutol susceptibility, however, when companion drug resistance was present in our study, the risk of acquiring rifampicin resistance was highest. For these patients, comprehensive drug susceptibility testing at isoniazid resistance detection may be necessary to prevent further resistance amplification.

Our study has several limitations. Twenty percent of collected specimens failed to yield viable cultures, although isoniazid resistance status was unknown at the time of collection, making differential attrition by exposure unlikely and limiting the risk of selection bias. Disease severity and bacillary burden were unmeasured but are more plausibly mediators than confounders, since samples were obtained at diagnosis before any differential treatment effect of undetected isoniazid resistance could influence disease severity. Residual confounding remains possible but would need to be extreme to explain the observed effect. Our transmission model applied the PAF as a fixed modifier and could therefore not capture the dynamic interaction between reduced acquired resistance and reductions in HR-TB transmission, likely making our projections conservative. Finally, the high prevalence of HR-TB and Lineage 2 in Vietnam means the PAF may overestimate the contribution of isoniazid resistance elsewhere, though the biological mechanism should hold broadly, as confirmed by our sensitivity analyses.

Isoniazid resistance, overwhelmingly under-detected by most current diagnostic strategies, is a near-necessary precondition for *de novo* rifampicin resistance acquisition under modern programmatic conditions. Diagnostic platforms designed to detect, and thereby combat, RR-TB have no effect on stemming the acquisition of rifampicin resistance. Expanding drug susceptibility testing at the time of TB diagnosis, combined with effective treatment, could substantially reduce the burden of RR-TB.

## Contributors

TMW conceptualised the study with input from GET and NTTT. Data generation and curation was performed by RS, NHH, PTP, LKL, KL, NNH, NLQ, DDAT, NTLH, TLTNT, TTBT, VTNH, DTMH, NHL, NPH, NHT, TTKQ, VD, SN and LHV. Formal analysis and visualisation was performed by RS with input from TMW, JAW and MC. Funding was acquired by TMW. RS, JAW, MC and TMW contributed to methodology. Project administration was conducted by NHH, PTP, RS, TMW, DTMH, NHL, NPH, NHT and TTKQ . Study resources were provided by TMW, GET, SN and DC. TMW, JAW and MC provided supervision. RS wrote the original draft with all authors reviewing and editing. RS and TMW verified the underlying data. All authors had full access to all the data in the study and had the final responsibility for the decision to submit for publication.

## Data sharing

All newly generated sequences are available in the European Nucleotide Archive under study accession PRJEB107150.

## Data Availability

All newly generated sequences are available in the European Nucleotide Archive under study accession PRJEB107150. Other data are available upon reasonable request to the authors.

## Acknowledgements

This research was funded in whole, or in part, by the Wellcome Trust (214560/Z/18/Z). RS is supported by the Rhodes Trust. TMW is a Wellcome Trust Clinical Career Development Fellow (214560/Z/18/Z).

## Declaration of generative AI and AI-assisted technologies in the manuscript preparation process

During the preparation of this work the author(s) used Claude AI in order to improve language and readability. After using this tool, the author(s) reviewed and edited the content as needed and take(s) full responsibility for the content of the published article.

## References

1. World Health Organization. Global Tuberculosis Report 2025. (2025).

2. World Health Organization. Automated Real-Time Nucleic Acid Amplification Technology for Rapid and Simultaneous Detection of Tuberculosis and Rifampicin Resistance: Xpert MTB/RIF System: Policy Statement. (2011).

3. Dean, A. S. et al. Prevalence and genetic profiles of isoniazid resistance in tuberculosis patients: A multicountry analysis of cross-sectional data. PLoS Med 17, e1003008 (2020).

4. Manson, A. L. et al. Genomic analysis of globally diverse Mycobacterium tuberculosis strains provides insights into the emergence and spread of multidrug resistance. Nat Genet 49, 395–402 (2017).

5. Gegia, M., Winters, N., Benedetti, A., Soolingen, D. van & Menzies, D. Treatment of isoniazid-resistant tuberculosis with first-line drugs: a systematic review and meta-analysis. The Lancet Infectious Diseases 17, 223–234 (2017).

6. Kendall, E. A., Fofana, M. O. & Dowdy, D. W. Burden of transmitted multidrug resistance in epidemics of tuberculosis: a transmission modelling analysis. The Lancet Respiratory Medicine 3, 963–972 (2015).

7. Spies, R., et al. Transmission of rifampicin-resistant tuberculosis in Ho Chi Minh City, Viet Nam: a prospective genomic epidemiology study. The Lancet Regional Health - Western Pacific 71, 101906 (2026).

8. Spies, R. et al. Spatial Analysis of Drug-Susceptible and Multidrug-Resistant Cases of Tuberculosis, Ho Chi Minh City, Vietnam, 2020–2023. Emerging Infectious Diseases 10.3201/eid3003.231309 (2024) doi:10.3201/eid3003.231309.

9. Bộ Y tế, Việt Nam (Ministry of Health). Hướng dẫn chẩn đoán, điều trị và dự phòng bệnh Lao [Guidelines for diagnosis, treatment and prevention of tuberculosis]. (2024).

10. Callum, J. et al. Prevalence and genetic basis of first-line drug resistance of Mycobacterium tuberculosis in Ca Mau, Vietnam. ERJ Open Res 8, 00122–02022 (2022).

11. Le Quang, N. et al. A modified decontamination and storage method for sputum from patients with tuberculosis. Wellcome Open Res 8, 166 (2023).

12. Westhead, J. et al. Characterizing the performance of an antibiotic resistance prediction tool, gnomonicus, using a diverse test set of 2,663 Mycobacterium tuberculosis samples. Microbial Genomics 11, 001592 (2025).

13. Walker, T. M. et al. Whole-genome sequencing to delineate Mycobacterium tuberculosis outbreaks: a retrospective observational study. The Lancet Infectious Diseases 13, 137–146 (2013).

14. World Health Organization. Catalogue of Mutations in Mycobacterium Tuberculosis Complex and Their Association with Drug Resistance. Second Edition. https://iris.who.int/bitstream/handle/10665/374061/9789240082410-eng.pdf?sequence=1 (2023).

15. Miettinen, O. S. PROPORTION OF DISEASE CAUSED OR PREVENTED BY A GIVEN EXPOSURE, TRAIT OR INTERVENTION1. Am J Epidemiol 99, 325–332 (1974).

16. Schwalb, A., et al. Potential impact, costs, and benefits of population-wide screening interventions for tuberculosis in Viet Nam: A mathematical modelling study. PLOS Global Public Health 5, e0005050 (2025).

17. Horton, K. C., Richards, A. S., Emery, J. C., Esmail, H. & Houben, R. M. G. J. Reevaluating progression and pathways following Mycobacterium tuberculosis infection within the spectrum of tuberculosis. Proceedings of the National Academy of Sciences 120, e2221186120 (2023).

18. Scarponi, D. et al. Demonstrating multi-country calibration of a tuberculosis model using new history matching and emulation package - *hmer*. Epidemics 43, 100678 (2023).

19. World Health Organization. WHO consolidated guidelines on tuberculosis. Module 4: Treatment and care. (2025).

20. Fregonese, F. et al. Comparison of different treatments for isoniazid-resistant tuberculosis: an individual patient data meta-analysis. The Lancet Respiratory Medicine 6, 265–275 (2018).

21. World Health Organization. Tuberculosis Profile: Viet Nam. https://worldhealthorg.shinyapps.io/tb_profiles/?_inputs_&entity_type=%22country%22&iso2=%22VN%22&lan=%22EN%22 (2025).

22. Levin, M. L. The occurrence of lung cancer in man. Acta Unio Int Contra Cancrum 9, 531–541 (1953).

23. Ford, C. B. et al. Mycobacterium tuberculosis mutation rate estimates from different lineages predict substantial differences in the emergence of drug-resistant tuberculosis. Nat Genet 45, 784–790 (2013).

24. Ektefaie, Y., Dixit, A., Freschi, L. & Farhat, M. R. Globally diverse Mycobacterium tuberculosis resistance acquisition: a retrospective geographical and temporal analysis of whole genome sequences. The Lancet Microbe 2, e96–e104 (2021).

25. Mitchison, D. A. Role of individual drugs in the chemotherapy of tuberculosis. The International Journal of Tuberculosis and Lung Disease 4, 796–806 (2000).

26. Kayigire, X. A., Friedrich, S. O., van der Merwe, L. & Diacon, A. H. Acquisition of Rifampin Resistance in Pulmonary Tuberculosis. Antimicrobial Agents and Chemotherapy 61, 10.1128/aac.02220-16 (2017).

27. World Health Organization. WHO Consolidated Guidelines on Tuberculosis. Module 3: Diagnosis. (2025).

28. Nguyen, T. M. et al. Molecular diagnostic tests for isoniazid-resistant tuberculosis: a scoping review. The Lancet Microbe 7, (2026).

29. Naidoo, K. & Dookie, N. Can the GeneXpert MTB/XDR deliver on the promise of expanded, near-patient tuberculosis drug-susceptibility testing? The Lancet Infectious Diseases 22, e121–e127 (2022).

30. Rudra, P. et al. A cartridge-based assay for improved detection of multidrug-resistant Mycobacterium tuberculosis directly from sputum. Journal of Clinical Microbiology 0, e01100–25 (2026).

