## Supplementary Methods for "Undetected isoniazid resistance leads to rifampicin-resistant tuberculosis: a prospective observational study and transmission modelling analysis"

**Supplementary appendix**

**Authors:**

Ruan Spies^a,b^, Nguyen Hong Hanh^a^, Phan Trieu Phu^a^, Luong Kim Lan^a^, Kim Lan^a^,
Ngo Ngoc Hue^a^, Nguyen Le Quang^a^, Do Dang Anh Thu^a^, Nguyen Thi Le Huong^a^,
Tran Le Thi Ngoc Thao^a^, Trinh Thi Bich Tram^a^, Vu Thi Ngoc Ha^a^, Dang Thi Minh Ha^c^, Nguyen Phuc Hai^c^, Nguyen Hung Thuan^c^, Tran Thi Kim Quy^c^, Nguyen Huu Lan^c^,
Viola Dreyer^d,e^, Stefan Niemann^d,e^, Derrick Crook^b,f^, Le Hong Van^a^, Guy Thwaites^a,b^,
Nguyen Thuy Thuong Thuong^a,b^, Marc Choisy^a,b^, James Watson^b^, Timothy Walker^a,b^

**Affiliations:**

^a^Oxford University Clinical Research Unit, Ho Chi Minh City, Viet Nam
^b^Nuffield Department of Medicine, University of Oxford, United Kingdom

^c^Pham Ngoc Thach Hospital, Ho Chi Minh City, Viet Nam

^d^German Center for Infection Research (DZIF), Partner Site Hamburg-Lübeck-Borstel-Riems, Borstel, Germany

^e^Research Center Borstel, Leibniz Lung Center, Molecular and Experimental Mycobacteriology, Borstel, Germany

^f^Shared Hospital Laboratory and Sunnybrook Research Institute, Sunnybrook Hospital, Toronto, Canada

Supplementary methods

Case-control study design
This study was originally designed as two nested case-control studies embedded within a prospective cohort of adults diagnosed with Xpert-confirmed RS-TB in Ho Chi Minh City between March 2020 and July 2024. We aimed to prospectively collect and archive sputum samples from all patients presenting with RS-TB at any of 23 DTUs or Phạm Ngọc Thạch Hospital. TB recurrence was defined as an Xpert-confirmed episode occurring at least 5 months after starting treatment for any prior episode.

The first case-control study aimed to investigate risk factors for RS-TB recurrence. Cases were defined as patients presenting with two Xpert-confirmed RS-TB episodes during the study period (n=400 target) and were enrolled consecutively as identified. No restriction on prior TB history was applied. Controls were randomly selected from patients with a single RS-TB episode at the same DTU as the corresponding case, matched by sex and age (±10 years), and were required to have had their first TB episode during the study period. Controls who developed recurrence during the study period were reclassified as cases and replaced.

**The second case-control study aimed to investigate** risk factors for acquired rifampicin resistance among patients with initially RS-TB. Cases were defined as patients whose index episode was Xpert RS-TB and whose recurrence episode was Xpert RR-TB (n=400 target), enrolled consecutively as identified. The 400 cases from the first case-control study (RS-TB recurrence) would serve as controls for the second.

Target sample sizes were calculated to provide at least 95% probability that the 95% confidence interval around estimated proportions of pre-existing isoniazid resistance in each group would fall within ±10 percentage points, with an allowance for attrition due to culture failure and reclassification. However, due to a low recruitment rate during the SARS-CoV-2 pandemic and later due to concerns about a high culture-negative rate among samples from recruited participants, we continued to recruit every possible recurrent case in the city throughout the duration of the study. Controls were recruited later in the study than planned because of the team’s workload who were also having to collect participant blood samples at different time-points for a parallel population-pharmacokinetic study (the subject of a subsequent manuscript).

During the study period, access to NTP notification data allowed estimation of population-level denominators for each outcome stratum: the number RS-TB notifications, RS-TB recurrences, and RR-TB recurrences. These denominators enabled the construction of sampling weights representing each participant's probability of selection into the analytical sample. This approach allowed direct estimation of risks, risk ratios, and population attributable fractions, avoiding the limitations of odds ratios from case-control studies, including non-collapsibility and limited interpretability. The primary exposure, outcome definitions, and analytical covariates remained unchanged from the original design.

Statistical analysis
Population denominators for each outcome stratum were derived from NTP notification data. During the study period, 42,843 patients were notified with RS-TB in Ho Chi Minh City, of whom 1,775 (4.1%) experienced at least one RS-TB recurrence. The RR-TB recurrence denominator was estimated at 550, extrapolated from a linked contemporaneous study in which 432 of 2,033 (21.2%) newly diagnosed RR-TB patients reported a prior RS-TB episode during the study period, applied to the 2,596 total RR-TB notifications during the study period.^1^ Sampling weights for each sub-cohort were the population-to-sample ratio. There were no missing covariate data.

Transmission model

Model structure
We developed a two-strain compartmental model of TB transmission, adapting previously described natural history frameworks^2,3^ to represent both DS and RR-TB. Model structure is presented in **Figure S2.** Model parameters and their prior ranges are provided in **Table S4**. The model was run from 1500 to 2024 with time-varying parameters for the care cascade and projected forward to 2035 under intervention scenarios. The model tracked a closed population of 100,000 individuals.

We represented TB natural history with six distinct states per strain.^4,5^ Model states are defined in **Table S5**. The model comprised 13 compartments in total (one shared Susceptible pool and six per strain). Disease state classification as infectious included both the Asymptomatic and Symptomatic compartments. Bidirectional transitions were permitted between Non-infectious and Asymptomatic states, and between Asymptomatic and Symptomatic states, reflecting the dynamic nature of TB disease progression and regression. Individuals in the Infection compartment could self-clear back to Susceptible, progress to Non-infectious disease, or progress directly to Asymptomatic TB. Non-infectious individuals could recover to Susceptible or progress to Asymptomatic TB. Diagnosis and treatment initiation occurred only from the Symptomatic compartment. Following treatment completion, individuals entered the Post-treatment compartment from which they could return to Susceptible or relapse to Asymptomatic disease.

##### Model equations

*Force of infection*

The strain-specific forces of infection are:

$$\lambda_{s}=\frac{\beta}{N}\left( \kappa\cdot A_{s}+C_{s} \right)$$

$$\lambda_{r}=\frac{\beta}{N}\cdot f\cdot\left( \kappa\cdot A_{r}+C_{r} \right)$$

where $\beta$ is the infectious contact rate, $N$ the population size, $A$ and $C$ the number of individuals with Asymptomatic and Symptomatic TB respectively, $\kappa$ the relative infectiousness of asymptomatic compared with symptomatic TB, and $f$ the relative fitness of the RR strain compared with the DS strain. Transmission occurs from both the Asymptomatic and Symptomatic compartments; all other compartments are non-infectious.

*Susceptible compartment*

$$\frac{\mathrm{dS}}{\mathrm{dt}}=\mu N+d_{s}C_{s}+d_{r}C_{r}+\eta\left( I_{s}+I_{r} \right)+\nu\left( N_{s}+N_{r} \right)+\tau\left( P_{s}+P_{r} \right)-\left( \lambda+\mu\right)S$$

Susceptible individuals are replenished by births ($\mu N$), TB-specific deaths ($d_{s}C_{s}+d_{r}C_{r}$), self-clearance of infection ($\eta$), recovery from non-infectious disease ($\nu$), and post-treatment return ($\tau$). They are depleted by infection (at total force $\lambda=\lambda_{s}+\lambda_{r}$) and background mortality ($\mu$).

*Drug-susceptible strain*

$$\frac{dI_{s}}{dt}=\lambda_{s}\cdot S-\left( \eta+p_{n}+p_{a}+\mu\right)\cdot I_{s}$$

$$\frac{dN_{s}}{dt}=p_{n}\cdot I_{s}+\zeta\cdot A_{s}-\left( \nu+\xi+\mu\right)\cdot N_{s}$$

$$\frac{dA_{s}}{dt}=p_{a}\cdot I_{s}+\xi\cdot N_{s}+\gamma\cdot C_{s}+\rho_{s}\left( 1-\alpha_{r} \right)\cdot P_{s}-\left( \zeta+\sigma+\mu\right)\cdot A_{s}$$

$$\frac{dC_{s}}{dt}=\sigma\cdot A_{s}+\phi_{s}\left( t \right)\left( 1-\alpha_{f} \right)\cdot T_{s}-\left( \gamma+\theta_{s}\left( t \right)+d_{s}\left( t \right)+\mu\right)\cdot C_{s}$$

$$\frac{dT_{s}}{dt}=\theta_{s}\left( t \right)\cdot C_{s}-\phi_{s}\left( t \right)\cdot T_{s}-\theta_{s}\left( t-\Delta_{s} \right)\cdot C_{s}\left( t-\Delta_{s} \right)\cdot e^{-\mu\Delta_{s}}-\mu\cdot T_{s}$$

$$\frac{dP_{s}}{dt}=\theta_{s}\left( t-\Delta_{s} \right)\cdot C_{s}\left( t-\Delta_{s} \right)\cdot e^{-\mu\Delta_{s}}-\left( \rho_{s}+\tau+\mu\right)\cdot P_{s}$$

From Infection ($I_{S}$), individuals may self-clear back to Susceptible ($\eta)$progress to Non-infectious disease ($p_{n}$), or progress directly to Asymptomatic TB ($p_{a}$). Bidirectional transitions occur between Non-infectious and Asymptomatic states (rates $\xi$ and $\zeta$) and between Asymptomatic and Symptomatic states (rates $\sigma$ and $\gamma$). Diagnosis and treatment initiation occur only from the Symptomatic compartment at rate $\theta_{S}\left( t \right)$. Treatment failure returns a fraction $\left( 1-\alpha_{f} \right)$ to Symptomatic DS-TB at rate $\phi_{S}\left( t \right)$. Treatment completion is modelled using a fixed-delay formulation: individuals who enter treatment at time $t-\Delta_{s}$ complete treatment at time $t$, attenuated by background mortality during the treatment period ($e^{-\mu\Delta_{s}}$), where $\Delta_{s}=0.5$ years corresponds to the 6-month DS-TB regimen. Following treatment, individuals either return to Susceptible ($\tau$) or relapse to Asymptomatic disease, with a fraction $\left( 1-\alpha_{r} \right)$ returning to Asymptomatic DS-TB at rate $\rho_{s}$.

*Resistance acquisition*

Two pathways transfer individuals from the DS to the RR strain:

$$\mathcal{A}_{f}=\phi_{s}\left( t \right)\cdot\alpha_{f}\cdot T_{s}$$

$$\mathcal{A}_{r}=\rho_{s}\cdot\alpha_{r}\cdot P_{s}$$

where $\mathcal{A}_{f}$represents resistance acquired through treatment failure (a fraction $\alpha_{f}$ of DS-TB treatment failures acquire rifampicin resistance, entering Symptomatic RR-TB directly) and $\mathcal{A}_{r}$ represents resistance acquired through post-treatment relapse (a fraction $\alpha_{r}$ of DS-TB relapses acquire resistance, entering Asymptomatic RR-TB).

*Rifampicin-resistant strain*

$$\frac{dI_{r}}{dt}=\lambda_{r}\cdot S-\left( \eta+p_{n}+p_{a}+\mu\right)\cdot I_{r}$$

$$\frac{dN_{r}}{dt}=p_{n}\cdot I_{r}+\zeta\cdot A_{r}-\left( \nu+\xi+\mu\right)\cdot N_{r}$$

$$\frac{dA_{r}}{dt}=p_{a}\cdot I_{r}+\xi\cdot N_{r}+\gamma\cdot C_{r}+\rho_{r}\cdot P_{r}+\mathcal{A}_{r}-\left( \zeta+\sigma+\mu\right)\cdot A_{r}$$

$$\frac{dC_{r}}{dt}=\sigma\cdot A_{r}+\phi_{r}\left( t \right)\cdot T_{r}+\mathcal{A}_{f}-\left( \gamma+\theta_{r}\left( t \right)+d_{r}\left( t \right)+\mu\right)\cdot C_{r}$$

$$\frac{dT_{r}}{dt}=\theta_{r}\left( t \right)\cdot C_{r}-\phi_{r}\left( t \right)\cdot T_{r}-\theta_{r}\left( t-\Delta_{r} \right)\cdot C_{r}\left( t-\Delta_{r} \right)\cdot e^{-\mu\Delta_{r}}-\mu\cdot T_{r}$$

$$\frac{dP_{r}}{dt}=\theta_{r}\left( t-\Delta_{r} \right)\cdot C_{r}\left( t-\Delta_{r} \right)\cdot e^{-\mu\Delta_{r}}-\left( \rho_{r}+\tau+\mu\right)\cdot P_{r}$$

The RR strain follows the same natural history structure as the DS strain, with two additions: the Asymptomatic compartment receives inflow from acquired resistance via relapse ($\mathcal{A}_{r})$ and the Symptomatic compartment receives inflow from acquired resistance via treatment failure ($\mathcal{A}_{f}$). Treatment failure in the RR strain returns individuals to Symptomatic RR-TB at rate $\phi_{R}\left( t \right)$ without further resistance change. Relapse from Post-treatment RR-TB returns individuals to Asymptomatic RR-TB at rate $\rho_{r}$.

*Incidence tracking*

To correctly attribute RR-TB incidence as transmitted or acquired without double-counting, we introduced a bookkeeping state variable $\hat{A}_{r}$ that tracks the subset of Asymptomatic RR-TB originating from relapse with acquired resistance. This variable follows the same outflow dynamics as $A_{r}$ but receives inflow only from $\mathcal{A}_{r}$:

$$\frac{d\hat{A}_{r}}{dt}=\mathcal{A}_{r}-\left( \zeta+\sigma+\mu\right)\cdot\hat{A}_{r}$$

This state does not affect transmission dynamics - the total $A_{r}$remains the full infectious pool for computing the force of infection. It is used solely to partition RR-TB incidence into its transmitted and acquired components:

$$\text{Inc}_{RR}^{tot}=\sigma\cdot A_{r}+\mathcal{A}_{f}$$

$$\text{Inc}_{RR}^{acq}=\sigma\cdot\hat{A}_{r}+\mathcal{A}_{f}$$

$$\text{Inc}_{RR}^{trans}=\sigma\cdot\left( A_{r}-\hat{A}_{r} \right)$$

where $Inc_{RR}^{total}$ is total RR-TB incidence (all new symptomatic RR-TB cases), $Inc_{RR}^{acq}$ is the component arising from acquired resistance, and $Inc_{RR}^{trans}$ is the component arising from transmission of pre-existing RR-TB strains. Acquired RR-TB incidence includes both the treatment failure pathway ($\mathcal{A}_{f}$, which enters Symptomatic RR-TB directly) and the relapse pathway (tracked via $\hat{A}_{r}$, which enters Symptomatic RR-TB upon progression at rate $\sigma$. The proportion of RR-TB attributable to acquired resistance is:

$$pAc=\frac{Inc_{RR}^{acq}}{Inc_{RR}^{total}}$$

DS-TB incidence is defined as $Inc_{DS}=\sigma\cdot A_{s}$

*Time-varying parameters*

Six care cascade parameters were modelled as piecewise linear forcing functions interpolating from an initial value (constant from 1900 to 1999) to a final value (reached by 2024), reflecting historical improvements in TB control in Vietnam:

$$p\left( t \right)=\left\{ \begin{matrix} p_{0} & \text{if }t\leq1999 \\ p_{0}+\left( p_{1}-p_{0} \right)\cdot\frac{t-1999}{25} & \text{if }1999<t<2024 \\ p_{1} & \text{if }t\geq2024 \end{matrix} \right.$$

This functional form was applied to the treatment initiation rate ($\theta$), treatment failure rate ($\phi$), and TB-specific mortality rate ($d)$ each separately for the DS and RR strains, giving 12 calibrated parameters (6 initial values and 6 final values; **Table S4**).

The RR-TB treatment duration ($\Delta_{r}$) followed a separate trajectory reflecting the rollout of shorter treatment regimens in Vietnam:

$$\Delta_{r}\left( t \right)=\left\{ \begin{matrix} 1.5 & \text{if }t\leq2020 \\ 1.5-1.0\cdot\frac{t-2020}{4} & \text{if }2020<t<2024 \\ 0.5 & \text{if }t\geq2024 \end{matrix} \right.$$

corresponding to a transition from an 18-month regimen ($\Delta_{r}=1.5$ years) to a 6-month BPaLM-based regimen ($\Delta_{r}=0.5$ years). The DS-TB treatment duration was fixed at $\Delta_{s}=0.5$ years throughout. After 2024, all time-varying parameters were held constant at their final values. Scenario projections therefore assume no further improvements in the care cascade beyond 2024, with the only change being the effect of the intervention on resistance acquisition rates.

Model calibration
The model was calibrated using history matching with emulation, implemented via the *hmer* R package.^6,7^ Twenty-eight parameters were estimated, with prior ranges informed by published natural history estimates **(Table S4)**. The model was simultaneously calibrated to 13 targets: WHO estimates of total TB incidence per 100,000 at seven time points (2000, 2005, 2010, 2015, 2019, 2023, 2024), RR-TB incidence per 100,000 at five time points (2015, 2017, 2019, 2023, 2024)^8^, and the proportion of RR-TB attributable to acquired resistance in 2024 (13–28%).^1^

Calibration proceeded iteratively. In the first wave, 560 parameter sets were sampled from the prior ranges using a maximin Latin hypercube design. Each was evaluated through the full delay differential equation model (solved using the dede function in the deSolve R package^9^), and a separate Gaussian process emulator was trained for each of the 13 calibration targets, yielding 13 emulators per wave. Emulators were validated using classification and comparison diagnostics; overconfident emulators had their variance inflated and poorly performing emulators were removed. Validated emulators were used to generate candidate parameter sets for the next wave. In subsequent waves, newly trained emulators were combined with those from the preceding waves, progressively refining the non-implausible region of parameter space. This process was repeated until 1,000 non-implausible parameter sets were generated. A stationarity analysis, in which the median intervention impact estimate was computed across repeated random subsamples of increasing size, confirmed that the estimate had stabilised, with the standard deviation across subsamples falling below 1 percentage point. Calibration diagnostics are presented in **Figure S4**.

Intervention mechanisms
Under intervention scenarios beginning in 2025, the resistance acquisition parameters $\alpha_{f}$ and $\alpha_{r}$ were reduced by a multiplicative factor reflecting the impact of isoniazid resistance diagnosis and treatment:

$$\alpha_{f}^{int}=\alpha_{f}\cdot\left( 1-c\cdot e\cdot PAF \right)$$

$$\alpha_{r}^{int}=\alpha_{r}\cdot\left( 1-c\cdot e\cdot PAF \right)$$

where $c$ is diagnostic coverage (the proportion of RS-TB patients tested for isoniazid resistance), $e$ is treatment efficacy (fixed at 0.89, corresponding to the estimated 1^st^ line treatment success rate in Vietnam^10^), and $PAF$ is the population attributable fraction of acquired RR-TB due to pre-existing isoniazid resistance. For each model run the $PAF$was sampled from a uniform distribution around its 95% confidence interval. Four coverage scenarios were evaluated: 20%, 50%, 80% and 100%.

Sensitivity analyses
To assess the robustness of our findings and to approximate the potential impact of isoniazid resistance diagnosis and treatment in settings with different isoniazid resistance prevalence or acquired resistance dynamics, we conducted a structured sensitivity analysis in which the PAF was recalculated across nine scenarios.

For the primary analysis, the PAF was estimated empirically using Miettinen’s formula, applying the prevalence of isoniazid resistance among participants who acquired RR-TB (cases) and the derived adjusted risk ratio ^11^:

$$PAF=\frac{p_{c}\times\left( RR-1 \right)}{RR}$$

where $p_{c}$is the prevalence of isoniazid resistance among participants who acquired rifampicin resistance (cases) and $RR$ is the adjusted risk ratio for acquired rifampicin resistance associated with pre-existing isoniazid resistance.

For the sensitivity analysis, we instead used Levin’s formula^12^:

$$PAF=\frac{p_{a}\times\left( RR-1 \right)}{1+p_{a}\times\left( RR-1 \right)}$$

where $p_{a}$ is the prevalence of isoniazid resistance among all TB patients. This formulation allows the PAF to be estimated from two quantities that are available for many settings globally: $p_{a}$ from drug resistance surveys and $RR$ from our empirical analysis, under the assumption that the biological mechanism linking isoniazid resistance to rifampicin resistance acquisition is conserved across settings.

Nine scenarios were defined by the full factorial combination of three levels of isoniazid resistance prevalence (low: 5%, average: 10%, high: 20%) and three levels of the relative risk for acquired rifampicin resistance (the empirically observed estimate, and 2-fold and 5-fold attenuations thereof). Uncertainty intervals for each $p_{a}$ and $RR$ combination were derived by Monte Carlo sampling (50,000 draws) from uniform distributions over plausible ranges for each parameter. The universal coverage intervention was then re-run under each of the nine PAF scenarios using the same parameter sets, with all other model parameters unchanged.

### Supplementary results

Risk of same-strain RS-TB recurrence by isoniazid resistance status
To examine whether pre-existing isoniazid resistance increased the risk of same-strain RS-TB recurrence, we restricted the RS-TB recurrence sub-cohort to participants with WGS data from both the index and recurrence episodes (n=150 of 422), as paired data were required to classify recurrences as same strain (≤12 SNPs) or reinfection (>12 SNPs). For the primary analysis, participants in the RS-TB recurrence sub-cohort required only an index isolate, since rifampicin susceptibility at recurrence was confirmed by Xpert. Covariate distributions, including isoniazid resistance prevalence, did not differ between those with and without paired data **(Table S6)**. Population denominators for all sub-cohorts were unchanged. Sampling weights for the RS-TB recurrence sub-cohort were recalculated to reflect the smaller sample size and sampling weights for the no recurrence sub-cohort were unchanged.

Among 150 participants with RS-TB recurrence and paired WGS data 107 (71%) had the same strain at recurrence and 43 (29%) were reinfections. The weighted risk of RS-TB relapse was 3.2% (95% CI 1.6–5.3) among participants with pre-existing isoniazid resistance and 3.1% (2.4–3.8) among those with isoniazid-susceptible TB, aRR 1.0 (0.5–1.8).

### Supplementary tables

#### Table S1: Isoniazid resistance-conferring mutations among index isolates, stratified by outcome

| Gene and mutation | Acquired rifampicin resistance  (N=49) | No acquired rifampicin resistance (N=151) |
| --- | --- | --- |
| *katG* S315T | 47 (96%) | 118 (78%) |
| *inhA* -777C>T | 4 (8%) | 34 (23%) |
| inhA S94A | 0 | 2 (1%) |
| *inhA* -770T>C | 0 | 2 (1%) |
| katG S315N | 0 | 1 (<1%) |

*Some isolates have more than 1 isoniazid resistance-conferring mutation

#### Table S2: Rifampicin resistance-conferring mutations among isolates with *de novo* acquired resistance

| Gene and mutation | N |
| --- | --- |
| *rpoB* H445R | 13 (25%) |
| *rpoB* H445Y | 10 (20%) |
| *rpoB* D435V | 9 (18%) |
| *rpoB* S450L | 8 (16%) |
| *rpoB* D435Y | 4 (8%) |
| *rpoB* H445D | 4 (8%) |
| rpoB L453P | 3 (6%) |
| *rpoB* Q432K | 3 (6%) |
| *rpoB* H445C | 2 (4%) |
| rpoB H445L | 2 (4%) |
| *rpoB* H445P | 1 (2%) |
| *rpoB* K446Q | 1 (2%) |
| *rpoB* L430P | 1 (2%) |
| *rpoB* Q432L | 1 (2%) |
| *rpoB* S450W | 1 (2%) |
| *rpoB* V170F | 1 (2%) |
| *rpoB* 1275_del_cggcac | 1 (2%) |
| *rpoB* 1291_ins_gcc | 1 (2%) |

*Some isolates have more than 1 rifampicin resistance-conferring mutation

#### Table S3: Risk of acquired rifampicin resistance by companion drug resistance pattern among participants with isoniazid-resistant TB

| **Resistance profile** | **n** | **Events** | **Weighted risk % (95% CI)** |
| --- | --- | --- | --- |
| Isoniazid only | 148 | 23 | 1.98 (1.23-2.94) |
| Isoniazid + pyrazinamide | 29 | 13 | 9.72 (4.14-30.58) |
| Isoniazid + ethambutol | 13 | 9 | 26.65 (7.26-94.91) |
| Isoniazid + pyrazinamide + ethambutol | 10 | 4 | 12.92 (1.82-71.41) |
| Isoniazid + any additional drug | 52 | 26 | 13.10 (6.95-31.26) |

#### Table S4: Model parameter descriptions, ranges and sources

| **Symbol** | **Description** | **Unit** | **Ranges** | **Source** |
| --- | --- | --- | --- | --- |
| $\beta$ | Infectious contact rate | capita^-1^yr^-1^ | 6.00-20.00 | ^2^ |
| $\kappa$ | Relative infectiousness from Asymptomatic TB compared to Symptomatic TB | - | 0.62-1.00 | ^13^ |
| $f$ | Relative fitness of RR strain compared to DS strain | - | 0.3-1 | ^14^ |
| $\eta$ | Rate of clearance from Infection back to Susceptible | capita^-1^yr^-1^ | 0.93-3.30 | ^3^ |
| $p_{n}$ | Rate of progression from Infection to Non-infectious TB | capita^-1^yr^-1^ | 0.04-0.23 | ^3^ |
| $p_{a}$ | Rate of progression from Infection to Asymptomatic TB | capita^-1^yr^-1^ | 0.01-0.1 | ^3^ |
| $\nu$ | Rate of recovery from Non-infectious TB back to Susceptible | capita^-1^yr^-1^ | 0.14-0.23 | ^3^ |
| $\xi$ | Rate of progression from Non-infectious TB to Asymptomatic TB | capita^-1^yr^-1^ | 0.21-0.28 | ^3^ |
| $\zeta$ | Rate of recovery from Asymptomatic TB to Non-infectious TB | capita^-1^yr^-1^ | 1.24-2.03 | ^3^ |
| $\sigma$ | Rate of progression from Asymptomatic TB to Symptomatic TB | capita^-1^yr^-1^ | 0.56-0.94 | ^3^ |
| $\gamma$ | Rate of recovery from Symptomatic TB back to Asymptomatic TB | capita^-1^yr^-1^ | 0.46-0.72 | ^3^ |
| $\theta_{s,0}$ | Rate of DS-TB treatment initiation from Symptomatic TB (initial) | capita^-1^yr^-1^ | 0.00-0.57 | ^2^ |
| $\theta_{s,1}$ | Rate of DS-TB treatment initiation from Symptomatic TB (final) | capita^-1^yr^-1^ | 0.57-0.77 | ^8^ |
| $\phi_{s,0}$ | Rate of DS-TB treatment failure (initial) | capita^-1^yr^-1^ | 0.11-1.00 | ^2^ |
| $\phi_{s,1}$ | Rate of DS-TB treatment failure (final) | capita^-1^yr^-1^ | 0.07-0.11 | ^8^ |
| $d_{s,0}$ | DS-TB specific mortality rate (initial) | capita^-1^yr^-1^ | 0.28-0.38 | ^15^ |
| $d_{s,1}$ | DS-TB specific mortality rate (final) | capita^-1^yr^-1^ | 0.00-0.28 | ^2^ |
| $\theta_{r,0}$ | Rate of RR-TB treatment initiation from Symptomatic TB (initial) | capita^-1^yr^-1^ | 0.00-0.30 | - |
| $\theta_{r,1}$ | Rate of RR-TB treatment initiation from Symptomatic TB (final) | capita^-1^yr^-1^ | 0.50-0.70 | - |
| $\phi_{r,0}$ | Rate of RR-TB treatment failure (initial) | capita^-1^yr^-1^ | 0.29-1.00 | - |
| $\phi_{r,1}$ | Rate of RR-TB treatment failure (final) | capita^-1^yr^-1^ | 0.07-0.29 | ^8^ |
| $d_{r,0}$ | RR-TB specific mortality rate (initial) | capita^-1^yr^-1^ | 0.35-0.50 | - |
| $d_{r,1}$ | RR-TB specific mortality rate (final) | capita^-1^yr^-1^ | 0.15-0.30 | ^8^ |
| $\tau$ | Rate of return from Post-treatment to Susceptible | capita^-1^yr^-1^ | 0.50-1.50 | ^16^ |
| $\rho_{s}$ | Rate of relapse from DS-TB Post-treatment | capita^-1^yr^-1^ | 0.01-0.3 | ^17^ |
| $\rho_{r}$ | Rate of relapse from RR-TB Post-treatment | capita^-1^yr^-1^ | 0.01-0.3 | - |
| $\alpha_{f}$ | Probability of acquiring rifampicin resistance during DS-TB treatment failure | - | 0.01-0.2 | - |
| $\alpha_{r}$ | Probability of acquiring rifampicin resistance during DS-TB relapse | - | 0.01-0.2 | - |
| $\mu$ | Background mortality rate per year (75-year life expectancy) | capita^-1^yr^-1^ | 0.013 | ^18^ |
| $\Delta_{s}$ | DS-TB treatment duration (6-month regimen) | years | 0.5 | - |
| $\Delta_{r,0}$ | RR-TB treatment duration (18-month regimen) | years | 1.5 | - |
| $\Delta_{r,1}$ | RR-TB treatment duration (6-month regimen) | years | 0.5 | - |

#### Table S5: Model compartment definitions

| **Symbol** | **State** | ***M. tuberculosis* bacilli** | **Inflammatory pathology** | **Symptoms** | **Infectious** | **Detectable by current diagnostic approach (passive case-finding)** | **Receiving anti-TB therapy** |
| --- | --- | --- | --- | --- | --- | --- | --- |
| $S$ | Susceptible | Absent | No | No | No | - | No |
| $I_{x}$ | *M. tuberculosis* infection | Present | No | No | No | No | No |
| $N_{x}$ | Non-infectious TB | Present | Yes | No | No | No | No |
| $A_{x}$ | Asymptomatic TB | Present | Yes | No | Yes (reduced) | No | No |
| $C_{x}$ | Symptomatic TB | Present | Yes | Yes | Yes | Yes | No |
| $T_{x}$ | Treatment | Present | Yes | Yes | No | - | Yes |
| $P_{x}$ | Post-treatment | Transition from present to absent | Resolving | No | No | - | No |

Subscript $x\in\{s,r\}$ denotes the drug-susceptible and rifampicin-resistant strain, respectively. The total population is $N=S+\sum_{x\in\{s,r\}} \left( I_{x}+N_{x}+A_{x}+C_{x}+T_{x}+P_{x} \right)$

#### Table S6: Characteristics of rifampicin-susceptible tuberculosis recurrence participants with and without paired whole genome sequencing data

|  | **Paired WGS available (N=150)** | **No paired WGS (N=272)** |
| --- | --- | --- |
| **Age, median (IQR)** | 53 (43-61) | 55 (46-61) |
| **Male sex, n** | 123 (82.0%) | 214 (78.7%) |
| **Diabetes, n** | 49 (32.7%) | 87 (32.0%) |
| **HIV, n** | 2 (1.3%) | 3 (1.1%) |
| ***M. tuberculosis* lineage, n** |  |  |
| Lineage 1 | 14 (9.3%) | 37 (13.6%) |
| Lineage 2 | 119 (79.3%) | 200 (73.5%) |
| Lineage 4 | 7 (4.7%) | 14 (5.1%) |
| Other/mixed | 10 (6.7%) | 21 (7.7%) |
| **Pre-existing isoniazid resistance, n** | 28 (18.7%) | 53 (19.5%) |
| **Pre-existing pyrazinamide resistance, n** | 3 (2.0%) | 9 (3.3%) |
| **Pre-existing ethambutol resistance, n** | 0 (0%) | 6 (2.2%) |

### Supplementary figures

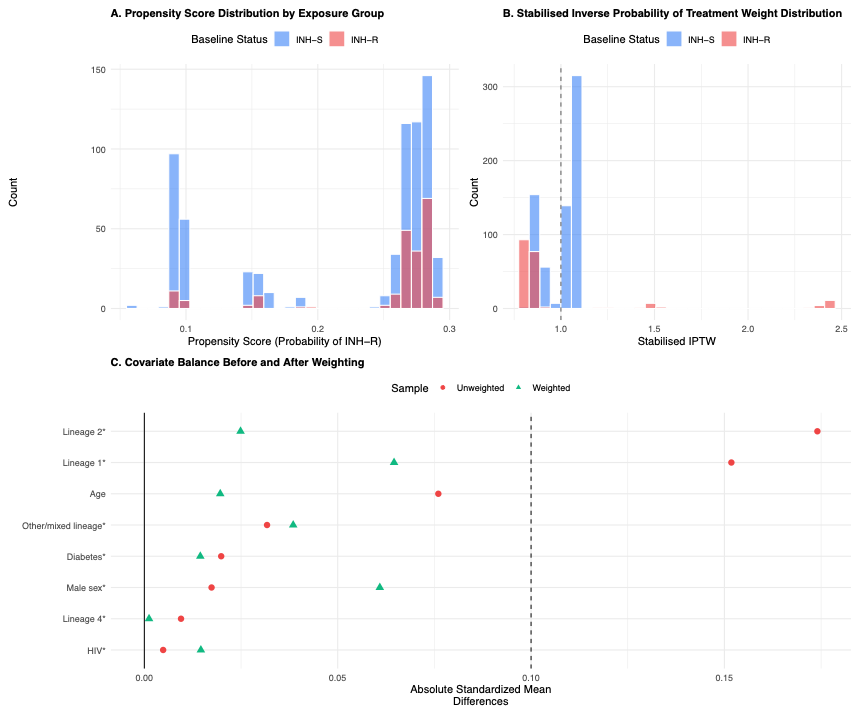

Figure S1: Inverse probability of treatment weighting diagnostics
**(A)** Propensity score distributions showing overlap between isoniazid-resistant (INH-R) and isoniazid-susceptible (INH-S) groups. **(B)** Distribution of stabilised inverse probability of treatment weights after truncation at the 1st and 99th percentiles; dashed line indicates a weight of 1 (no reweighting). **(C)** Absolute standardised mean differences for all covariates before (unweighted) and after weighting; dashed line indicates the 0.1 threshold for adequate balance.

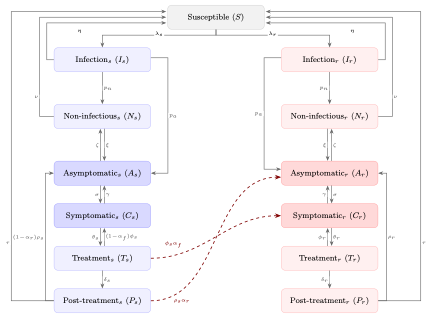

Figure S2: Two-strain compartmental model of tuberculosis transmission and rifampicin resistance acquisition.
Schematic of the deterministic compartmental model representing drug-susceptible (S, blue) and rifampicin-resistant (R, pink) tuberculosis. The model comprises 13 compartments: one shared Susceptible pool and six disease states per strain (Infection, Non-infectious, Asymptomatic, Symptomatic, Treatment, Post-treatment). Solid arrows indicate within-strain transitions; dashed red arrows indicate cross-strain resistance acquisition pathways, whereby a fraction of drug-susceptible treatment failures and post-treatment relapses acquire rifampicin resistance and enter the rifampicin-resistant strain.

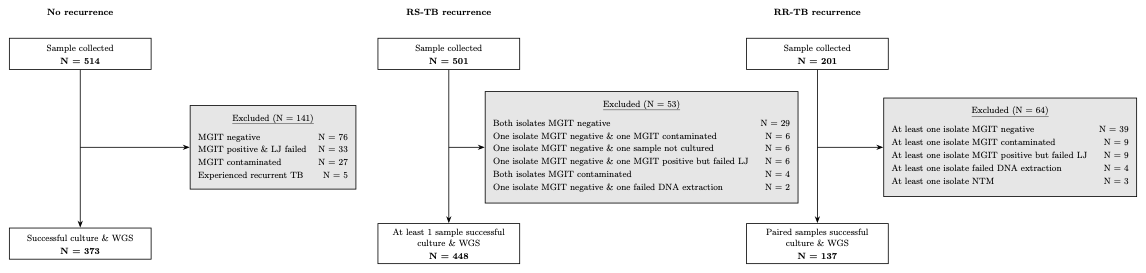

#### Figure S3: Reasons for exclusion between sample collection and successful whole genome sequencing

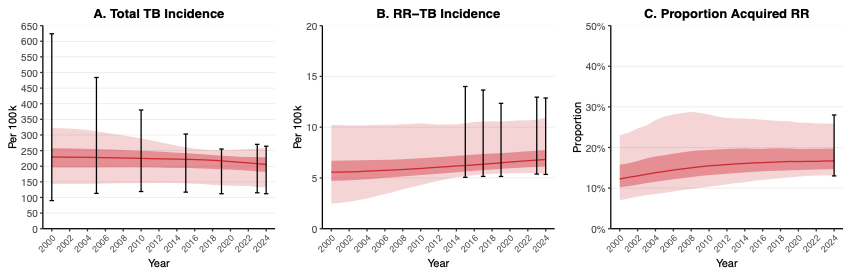

Figure S4: Model calibration to observed epidemiological data for Vietnam, 2000-2024
Model output trajectories (red) compared with calibration targets (black error bars) for total TB incidence (A), rifampicin-resistant TB incidence (B), and the proportion of RR-TB attributable to acquired resistance (C). Solid lines represent medians, dark shaded areas the interquartile range, and light shaded areas the 95% uncertainty interval across 1,000 non-implausible parameter sets identified by history matching with emulation

### References

1.	Spies, R. *et al.* Transmission of rifampicin-resistant tuberculosis in Ho Chi Minh City, Viet Nam: a prospective genomic epidemiology study. 2026.03.21.26348963 Preprint at https://doi.org/10.64898/2026.03.21.26348963 (2026).

2.	Schwalb, A. *et al.* Potential impact, costs, and benefits of population-wide screening interventions for tuberculosis in Viet Nam: A mathematical modelling study. *PLOS Global Public Health* **5**, e0005050 (2025).

3.	Horton, K. C., Richards, A. S., Emery, J. C., Esmail, H. & Houben, R. M. G. J. Reevaluating progression and pathways following Mycobacterium tuberculosis infection within the spectrum of tuberculosis. *Proceedings of the National Academy of Sciences* **120**, e2221186120 (2023).

4.	Coussens, A. K. *et al.* Classification of early tuberculosis states to guide research for improved care and prevention: an international Delphi consensus exercise. *The Lancet Respiratory Medicine* **12**, 484–498 (2024).

5.	World Health Organization. *Global Tuberculosis Report 2024*. (2024).

6.	Iskauskas, A. *et al.* Emulation and History Matching Using the hmer Package. *Journal of Statistical Software* **109**, 1–48 (2024).

7.	Scarponi, D. *et al.* Demonstrating multi-country calibration of a tuberculosis model using new history matching and emulation package - *hmer*. *Epidemics* **43**, 100678 (2023).

8.	World Health Organization. *Global Tuberculosis Report 2025*. (2025).

9.	Soetaert, K., Petzoldt, T. & Setzer, R. W. Solving Differential Equations in R: Package deSolve. *Journal of Statistical Software* **33**, 1–25 (2010).

10.	World Health Organization. *Tuberculosis Profile: Viet Nam*. https://worldhealthorg.shinyapps.io/tb_profiles/?_inputs_&entity_type=%22country%22&iso2=%22VN%22&lan=%22EN%22 (2025).

11.	MIETTINEN, O. S. PROPORTION OF DISEASE CAUSED OR PREVENTED BY A GIVEN EXPOSURE, TRAIT OR INTERVENTION1. *Am J Epidemiol* **99**, 325–332 (1974).

12.	Levin, M. L. The occurrence of lung cancer in man. *Acta Unio Int Contra Cancrum* **9**, 531–541 (1953).

13.	Emery, J. C. *et al.* Estimating the contribution of subclinical tuberculosis disease to transmission: An individual patient data analysis from prevalence surveys. *eLife* **12**, e82469 (2023).

14.	Luciani, F., Sisson, S. A., Jiang, H., Francis, A. R. & Tanaka, M. M. The epidemiological fitness cost of drug resistance in Mycobacterium tuberculosis. *Proceedings of the National Academy of Sciences* **106**, 14711–14715 (2009).

15.	Richards, A. S. *et al.* Quantifying progression and regression across the spectrum of pulmonary tuberculosis: a data synthesis study. *The Lancet Global Health* **11**, e684–e692 (2023).

16.	Ruan, Q. *et al.* Recurrent pulmonary tuberculosis after treatment success: a population-based retrospective study in China. *Clinical Microbiology and Infection* **28**, 684–689 (2022).

17.	Huyen, M. N. T. *et al.* Tuberculosis Relapse in Vietnam Is Significantly Associated With Mycobacterium tuberculosis Beijing Genotype Infections. *J Infect Dis* **207**, 1516–1524 (2013).

18.	World Bank Open Data. *World Bank Open Data* https://data.worldbank.org.
